# Mobile applications for atrial fibrillation self-management: a systematic search and evaluation

**DOI:** 10.1101/2025.07.09.25331174

**Authors:** Wojciech Losos, Carrie Shao, Biqi Wang, Spyros Kitsiou, Ben S. Gerber, David D. McManus

**Author notes:** Corresponding author (WL). These authors contributed equally to this work (Wojciech Losos, Carrie Shao).

## Abstract

Atrial fibrillation (AF) is the most common cardiac arrhythmia, increasing the risk of stroke, heart failure, and healthcare costs. Although patient self-management can improve outcomes, sustaining long-term engagement is often difficult. Mobile health applications may help address this gap, but their quality and clinical alignment have not been systematically assessed using a validated framework.

A structured search of the Apple App Store and Google Play Store identified free, English-language apps supporting AF self-management. Eligible apps included features such as symptom tracking, medication reminders, or educational content. App quality was assessed using the Mobile Application Rating Scale (MARS), which evaluates engagement, functionality, aesthetics, and information quality.

Of 455 apps identified, five met all inclusion criteria. Common features included symptom tracking and medication logging, but coverage of evidence-based care domains varied. Mean MARS scores ranged from 4.07 to 4.53 out of 5. Higher-performing apps excelled in functionality and information quality but often lacked comprehensive integration of guideline-recommended care, such as stroke risk assessment or personalized feedback.

These findings highlight a gap in high-quality, clinically grounded digital tools for AF self-care. Improved co-design processes and clearer frameworks for app evaluation may help guide the development and selection of effective tools to support AF self-management.

**Author Summary:** We wanted to understand whether smartphone apps can help people manage atrial fibrillation (AF), a common heart rhythm condition that raises the risk of stroke and heart failure. While actively managing AF through lifestyle changes and regular monitoring can reduce health risks, many people find it hard to stay engaged over time. Mobile apps could make self-care more convenient, but their quality and usefulness vary greatly. We searched the two largest app stores for free apps in English that offer tools to help people track symptoms, manage medications, and learn about their condition. Out of more than 450 apps, only five met our criteria. We found that while these apps include helpful features, they often do not cover all aspects of AF care or follow current medical guidelines closely. Our findings show that people with AF have limited trustworthy options to support self-care. We hope our study encourages app developers, healthcare professionals, and patients to work together to create better digital tools that can safely and effectively support people living with AF.

## Introduction

Atrial fibrillation (AF) is the most prevalent cardiac arrhythmia globally, significantly increasing the risk of stroke, heart failure, and healthcare costs [1]. With an aging global population and a rising prevalence of chronic diseases, effective AF management has become a major public health priority [2]. Patient self-management, including medication adherence, symptom monitoring, rhythm tracking, and lifestyle behavior adjustments (e.g., heart healthy diet, physical activity, and smoking cessation), can play a critical role in controlling AF and improving clinical outcomes [3]. Notably, landmark studies such as LEGACY and LEAF have demonstrated the substantial benefits of sustained weight loss and comprehensive lifestyle modifications in reducing AF burden [4,5]. While structured education and clinical support can improve adherence and quality of life in the short term, maintaining long-term patient engagement remains a persistent challenge [3,6].

In response to these challenges, mobile health (mHealth) technologies—including smartphone applications integrated with wearable devices—offer the potential to support continuous rhythm monitoring, medication adherence, and lifestyle modification [7,8]. However, the quality and clinical utility of AF-focused apps remain highly variable. A prior review by Turchioe et al. examined mobile applications for AF detection and management, highlighting the diversity of available tools and the growing role of digital health in this space [9]. Their review included both patient- and clinician-facing apps and focused largely on rhythm detection capabilities, without evaluating app quality using a validated framework.

Although some prior work has evaluated AF-related apps, including some with management features, no study to date has systematically assessed patient-facing AF self-management applications using a validated tool such as the Mobile Application Rating Scale (MARS) or examined their alignment with evidence-based self-care practices. Our study addresses this gap by focusing specifically on tools designed to support patient engagement in AF self-management—such as symptom tracking, medication adherence, and lifestyle modification—and applying a structured rating scale to evaluate app quality across multiple domains.

Despite the growing number of digital tools, the evidence remains fragmented, and patients lack clear guidance on trustworthy, clinically grounded self-management options. Therefore, this study aimed to systematically identify, characterize, and evaluate patient-facing AF self-management apps using the validated Mobile Application Rating Scale (MARS), and to assess how well these apps align with evidence-based care practices.

## Results

### Search Results

A total of 455 apps were identified through systematic searches of the Apple App Store and Google Play. After removing duplicates and ineligible versions, 320 unique apps remained for title and description screening. Of these, 310 were excluded for not meeting inclusion criteria. Ten apps were downloaded for full eligibility assessment, and five met all criteria for final analysis. Four of these were identified through app store searches, and one was identified through our supplementary literature review. A summary of the selection process is provided in Fig 1.

**Fig 1.**
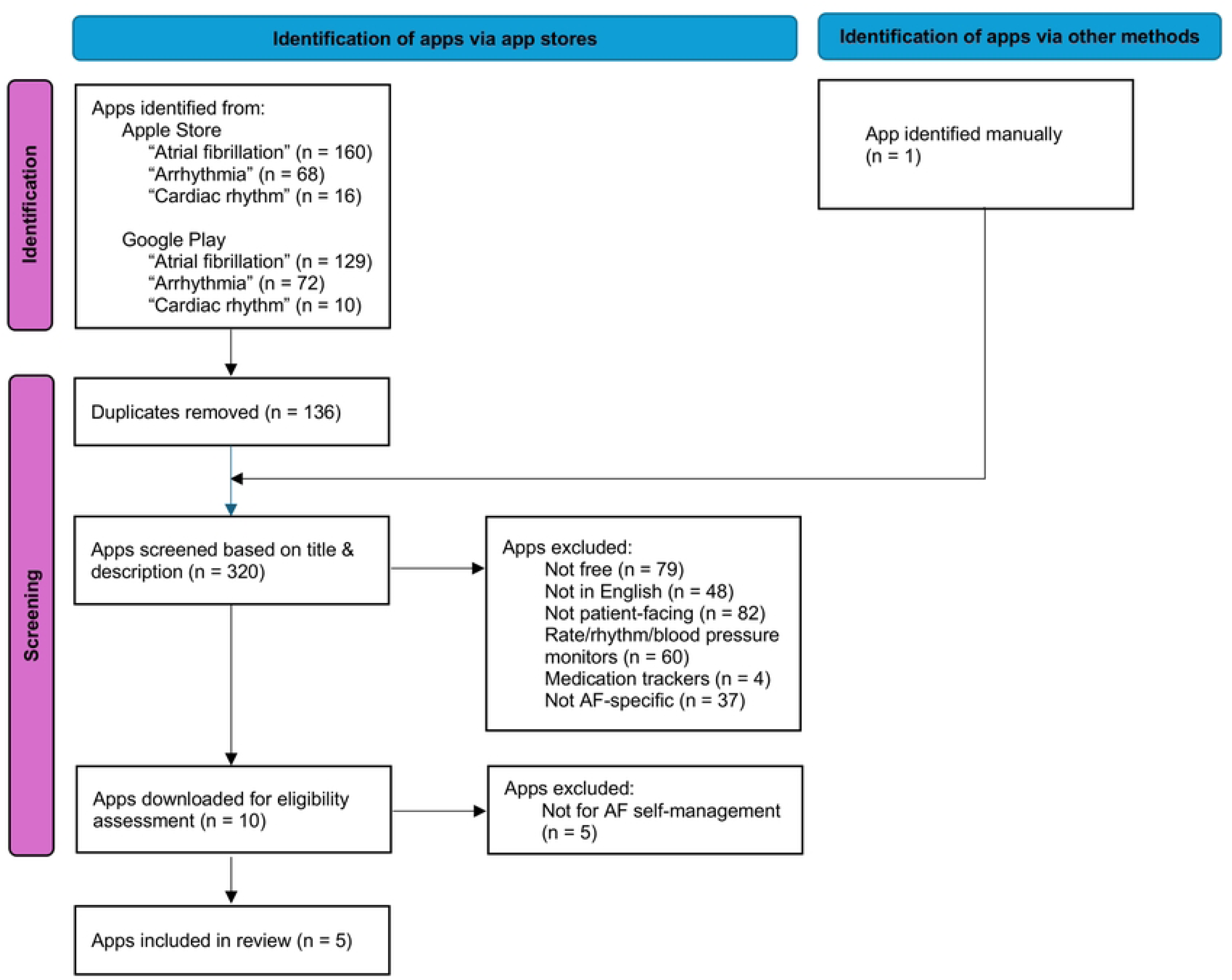
PRISMA flow diagram of app screening.

### App Characteristics and Functionalities

Key app characteristics are summarized in Table 1. All five apps are available on iOS, and four are also available on Android. While each is free to download, one (*AFibLife*) offers optional in-app purchases. Although not exclusively designed for AF, *Health Storylines* provides modular, customizable self-management tools—such as symptom tracking, medication logging, and a health diary—that can be adapted by patients with AF to monitor their condition and treatment adherence. Its inclusion highlights the practical relevance of general-purpose self-management apps within the AF digital health landscape, acknowledging how patients commonly use such tools alongside condition-specific apps.

**Table 1.** Characteristics of included AF self-management apps. Developer, platform, version, update date, cost, primary purpose, and major features are summarized.

| APP NAME | DEVELOPER | PLATFORM | VERSION | LAST UPDATE | COST | PRIMARY PURPOSE | DATA PRIVACY & SECURITY | CONNECTIVITY & INTEGRATION | LANGUAGES AVAILABLE | FEATURES & FUNCTIONALITY |
| --- | --- | --- | --- | --- | --- | --- | --- | --- | --- | --- |
| AFIB MANAGER | @Point of Care | iOS | 11.0.4 | Oct 26, 2023 | Free | AFib management, data integration with Health app | Not described in public listing | Integrates with Health app, compatible with heart rate devices | English | Heart rate data integration, AFib management tools |
| AFIB 2GETHER | Pfizer Inc. | iOS, Android | 1.0.0 | Jul 11, 2022 | Free | Education on AFib, stroke risk assessment, shared decision-making | Collects personal info from users and healthcare providers | Not specified | English | Stroke risk questionnaire, educational resources, FAQ tool |
| CLIEXA-PULSE | cliexa | iOS, Android | 1.0.0 | Jan 15, 2024 | Free | Symptom tracking, medication management, episode logging | Not described in public listing | Not specified | English | Symptom tracking, medication logging, episode documentation |
| AFIBLIFE | Appmanschap B.V. | iOS | 1.5.0 | Jan 23, 2025 | Free (In-App Purchases) | AFib education, quality of life assessment, health data integration | Data may be collected and linked to user identity | Option to connect health data from mobile device | English, German, Japanese, Portuguese | AFEQT questionnaire, health data integration, journaling, stroke risk calculator |
| HEALTH STORYLINES | Self Care Catalysts | iOS, Android | 10.0.0 | Oct 10, 2023 | Free | Symptom tracking, medication adherence, health diary, customizable care plans | Data collected and user-controlled sharing options | Synchs with Apple Health, Google Fit, some wearable devices | English | Symptom tracking, medication management, health education, daily journaling, customizable trackers |

Coverage of self-management domains varies across the apps (Table 2). *AFib Manager* offers comprehensive functionality, including symptom tracking, medication logging, episode recording, educational content, and health device integration. However, it does not include stroke risk assessment or shared decision-making support. *AFib 2gether* focuses on education, stroke risk communication, and decision support but lacks monitoring and tracking capabilities. *cliexa-PULSE* includes symptom tracking, medication management, and episode logging but does not provide educational content or stroke risk tools. *AFibLife* supports all evaluated domains, though its medication feature is limited to logging only and does not include medication reminders, refill alerts, or integration with pharmacy or adherence tracking tools. *Health Storylines* provides a customizable platform and medication tracking that, while not AF-specific, aligns with common self-care needs for AF patients and demonstrates the functional versatility patients may seek in general health apps.

**Table 2.** Coverage of core AF self-management domains across included apps. Domains include symptom tracking, medication management, episode logging, education, stroke risk assessment, shared decision-making, and device integration.

| APP NAME | SYMPTOM TRACKING | MEDICATION MANAGEMENT | EPISODE LOGGING | EDUCATION/ INFORMATION | STROKE RISK ASSESSMENT | SHARED DECISION- MAKING SUPPORT | DATA INTEGRATION WITH HEALTH DEVICES | TOTAL FUNCTIONS COVERED |
| --- | --- | --- | --- | --- | --- | --- | --- | --- |
| AFIBLIFE | Yes | Partial (Logging only) | Yes | Yes | Yes | Yes | Yes | 6.5 |
| AFIB MANAGER | Yes | Yes | Yes | Yes | No | No | Yes | 5 |
| HEALTH STORYLINES | Yes | Yes | No | Yes | No | No | Yes | 4 |
| CLIEXA-PULSE | Yes | Yes | Yes | No | No | No | No | 3 |
| AFIB 2GETHER | No | No | No | Yes | Yes | Yes | No | 3 |

*Note: Partial functionality (e.g., medication logging without reminders) was scored as 0.5.*

### App Quality Assessment

Consensus MARS scores ranged from 4.07 to 4.53 (Table 3). *AFib 2gether* and *AFibLife* scored highest (4.53), followed by *cliexa-PULSE* (4.38). *AFib 2gether* had the highest score in functionality (5.00) and shared the highest information quality score (4.83). *cliexa-PULSE* had the highest aesthetic score (5.00). *Health Storylines* received the highest engagement score (4.40) but scored lower in functionality (3.75) and information quality (4.17), resulting in a moderate overall score (4.08). *AFib Manager* had the lowest mean score (4.07), primarily due to a lower aesthetic rating (3.67). Overall, apps that performed well across multiple MARS domains— particularly functionality and information—had the highest quality scores.

**Table 3.** Mean Mobile Application Rating Scale (MARS) scores across four quality domains. Domains include engagement, functionality, aesthetics, and information quality.

| App | Engagement | Functionality | Aesthetics | Information | Mean App Quality |
| --- | --- | --- | --- | --- | --- |
| AFib Manager | 4.20 | 4.25 | 3.67 | 4.17 | 4.07 |
| AFib 2gether | 3.60 | 5.00 | 4.67 | 4.83 | 4.53 |
| cliexa-PULSE | 4.20 | 3.50 | 5.00 | 4.83 | 4.38 |
| AFibLife | 4.20 | 4.75 | 4.33 | 4.83 | 4.53 |
| Health Storylines | 4.40 | 3.75 | 4.00 | 4.17 | 4.08 |

*Higher scores reflect better app performance; each subdomain rated on a 1–5 scale.*

### Inter-Rater Reliability

Inter-rater agreement across apps was fair overall (Krippendorff’s alpha = 0.53). Reliability varied by app. *Health Storylines* showed the highest agreement (α = 0.69), while *AFib Manager* showed the lowest (α = 0.274). *Health Storylines* also had the smallest mean-score difference between raters (–0.088). Its modular design and clear labeling likely facilitated uniform interpretation. Apps with lower agreement often had less intuitive layouts or ambiguous content.

## Discussion

We found general acceptability but considerable variability in the quality and comprehensiveness of the five available AF self-management apps we reviewed. While all included core features— such as symptom tracking and medication logging—none offered a fully integrated, guideline-aligned platform encompassing rhythm monitoring, patient education, behavioral support, and shared decision-making tools. This highlights a critical gap in the availability of mHealth resources that comprehensively support AF self-management in line with evidence-based care. In particular, structured lifestyle interventions (e.g., weight management, alcohol moderation), individualized stroke risk education, and long-term adherence support were often absent or only partially addressed. Despite increasing interest in digital health, many AF apps still fall short of delivering clinically robust, user-centered functionality [3–5,9].

Our findings align with prior evaluations of cardiovascular mHealth tools, especially for heart failure self-management, which have similarly reported wide variation in quality, limited use of behavioral frameworks, and inconsistent alignment with clinical guidelines [11]. Many earlier reviews relied on narrative summaries or author-developed checklists, reducing reproducibility and comparability across studies. By contrast, our study used the validated Mobile Application Rating Scale (MARS) [10], providing a structured assessment across four domains— Engagement, Functionality, Aesthetics, and Information Quality—and incorporated inter-rater reliability to enhance objectivity. This dual focus enabled us to differentiate apps not only by their content but also by the clarity and consistency of their user interface—a dimension often overlooked in prior reviews [9].

Functionality and aesthetic scores varied widely, reflecting persistent challenges in app usability and design. These disparities likely stem from limited application of user-centered design (UCD) principles, such as intuitive navigation, visual clarity, and mobile responsiveness—factors known to affect both adoption and sustained use [10]. Superficial features alone are insufficient; meaningful engagement depends on seamless and accessible interfaces. Notably, even the highest-rated apps demonstrated limited personalization and dynamic feedback, mirroring broader trends in cardiovascular app development where tailored features remain underutilized despite their proven role in supporting behavior change, medication adherence, and self-efficacy [11,14].

Clear design and structural consistency are vital not only for usability but also for evaluability — key considerations for developers and researchers aiming to recommend digital tools [10]. Integrating adaptive elements, such as customizable goals, personalized alerts, and feedback loops informed by user behavior, may help drive deeper engagement. Researchers should carefully evaluate apps not only for content but also for design and functionality, as these dimensions directly impact patient engagement. Future research should explore how specific design features affect long-term adherence and whether embedding behavioral theory into app development can bridge the gap between initial interest and sustained use.

This study has several limitations. First, we restricted inclusion to free, English-language apps, which may have excluded high-quality paid or non-English options. For example, subscription-based apps such as *Kardia Premium* offer rhythm monitoring and patient education but were excluded due to cost or because they primarily focus on device integration rather than comprehensive self-management. The prevalence of such apps suggests our sample may not fully capture the broader mHealth landscape for AF. Additionally, we included *Health Storylines*, a general chronic disease self-management app, to reflect how patients often adapt multi-condition tools to support AF self-care alongside other comorbidities in real-world practice. Second, although MARS provides a structured and validated framework [10], it may not fully capture real-world clinical relevance or patient usability. Insights gained through patient feedback after extended use could reveal trust, engagement, and usability factors that structured appraisals alone may not detect. Third, because mobile apps are frequently updated, features and design elements may have changed since our evaluation. Fourth, we did not include direct clinician input in our app assessments, which could have added perspectives on content accuracy, workflow integration, and alignment with evidence-based care pathways [11]. Finally, our privacy and security assessment was limited to the information provided in app store descriptions and linked privacy policies; we did not conduct a detailed legal or technical audit of each app’s user agreement or data handling practices.

To ensure relevance, sustainability, and equity in AF self-management, future app development should prioritize interdisciplinary co-design involving clinicians, behavioral scientists, engineers, and patients. Co-design frameworks—such as participatory and human-centered design—can better align digital tools with user preferences and real-world needs [14]. Engaging patients early in development, through co-creation workshops or citizen science approaches, has been shown to improve usability, trust, and uptake [14,15]. These approaches are particularly important for older adults with AF, who may face barriers such as low digital literacy, interface complexity, and reduced confidence in using mobile tools [15]. They may also experience physical limitations (e.g., visual or motor impairments) and less prior exposure to technology—factors that further complicate engagement [16]. By integrating patient and clinician perspectives, developers can produce tools that are both clinically grounded and user-friendly, increasing the likelihood of sustained engagement in AF care [6,7]. As of 2020, AF affects more than 46 million people worldwide, with prevalence projected to rise as populations age [1,2]. Despite the growing number of mHealth tools, adoption among people with AF—particularly older adults—remains limited, highlighting the need for inclusive, user-informed design and implementation strategies [15].

In summary, our findings underscore the need for greater transparency, standardization, and oversight in the development and evaluation of AF self-management apps. In the absence of regulatory oversight, clinicians and patients must navigate a largely unregulated digital marketplace, increasing the risk of adopting tools with limited clinical accuracy or usability. Addressing this gap will require evidence-based frameworks and endorsement mechanisms— such as digital formularies or certification programs—to guide the safe integration of high-quality apps into routine care. Incorporating app assessments into shared decision-making, particularly for rhythm monitoring and lifestyle interventions, could further strengthen patient-centered care. At the policy level, developing public-facing repositories or app rating platforms that combine expert review and patient feedback may help bridge the divide between technological innovation and clinical utility in digital health. Future research should also examine how co-designed apps impact long-term patient engagement and clinical outcomes.

## Methods

### Search Strategy and App Identification

We systematically searched the Apple iOS Store and Google Play Store in June 2024 and updated the search in January 2025 using the terms: “atrial fibrillation,” “arrhythmia,” and “cardiac rhythm.” Apps were included if they met all of the following criteria: (1) free to download, (2) available in English, (3) designed for patient use, (4) explicitly referenced atrial fibrillation, and (5) offered at least one self-management feature such as symptom logging, medication reminders, or lifestyle education. In addition to apps specifically labeled for AF, we also included general self-management apps if they offered features that could plausibly support AF self-care in real-world use (e.g. symptom tracking, medication adherence, vital signs monitoring, education). This strategy aligns with evidence that patients frequently use multi-condition apps to manage AF alongside other chronic conditions [11]. To supplement the app store search and ensure comprehensive coverage, we reviewed prior systematic reviews of cardiovascular mHealth apps to identify additional apps relevant to AF self-care not captured by our keyword strategy.

When an app was available on both iOS and Android platforms, we retained the iOS version to ensure consistency in evaluation. Similar versions of the same app (e.g., “Pro” or “Lite” editions) were treated as duplicates, and only the most complete or commonly downloaded version was retained for screening. We excluded general heart rate or rhythm monitors without self-management features, blood pressure tracking apps, clinician-only platforms, and apps requiring institutional logins.

### App Characterization

For each included app, we extracted descriptive data, including app developer, version, release/update dates, platform availability, cost, main functions, supported languages, privacy disclosures, and wearable integration capabilities. Data privacy and security information was extracted from each app’s store description and the linked privacy policy when available; a full legal review of user agreements was not conducted.

### App Quality Assessment

App quality was evaluated using the Mobile Application Rating Scale (MARS) [10], which assesses four domains: Engagement (entertainment, interest, customization, interactivity, target group), Functionality (performance, ease of use, navigation, gestural design), Aesthetics (layout, graphics, visual appeal), and Information (accuracy of app description, goals, quality and quantity of information, visual information, credibility, evidence base). Each subcategory was rated on a 5-point Likert scale (1 = inadequate, 5 = excellent); domain scores were then averaged to calculate a composite mean quality score for each app.

### Reviewer Training and Reliability

Two independent reviewers completed a calibration session using three non-study apps to standardize MARS scoring before independently rating each app. Discrepancies were resolved through consensus to determine final scores. Inter-rater reliability was quantified using Krippendorff’s Alpha in R (version 4.3.3) [13], treating item scores as ordinal variables. Krippendorff’s Alpha is a robust reliability coefficient suitable for ordinal data and small sample sizes and has been widely adopted in health technology evaluation studies [12].

### Ethics Statement

This study did not involve human participants, and no identifiable personal data were collected or analyzed. Therefore, institutional review board approval and informed consent were not required. All app-level MARS scores and reviewer ratings are included in Table 3, and no additional data were excluded or withheld.

## Author Contributions

Conceptualization: W.L., C.S., D.D.M.;

Methodology: W.L., B.W., S.K.;

Data Curation: B.W., C.S.;

Formal Analysis: W.L., B.W.;

Investigation: W.L., C.S., B.W.;

Writing – original draft: W.L., C.S.;

Writing – review & editing: S.K., B.S.G., D.D.M.;

Supervision: S.K., B.S.G., D.D.M.;

Funding Acquisition: D.D.M.

## Data Availability

All data generated or analyzed during this study are included in this published article. There are no additional data.

## References

1. Xu L, Chen Y, Lin J, et al. Burden of atrial fibrillation and its attributable risk factors from 1990 to 2019: An analysis of the Global Burden of Disease study 2019. Lancet Reg Health West Pac. 2022;23:100402. doi: 10.1016/j.lanwpc.2022.100402.

2. Lip GYH. The ABC pathway: an integrated approach to improve AF management. Nat Rev Cardiol. 2017;14:627–628. doi: 10.1038/nrcardio.2017.153.

3. Desteghe L, Engelhard L, Vijgen J, et al. Effect of reinforced, targeted in-person education using the Jessa Atrial fibrillation Knowledge Questionnaire in patients with atrial fibrillation: A randomized controlled trial. Eur J Cardiovasc Nurs. 2019;18:194–203. doi: 10.1177/1474515118804353.

4. Pathak RK, Middeldorp ME, Meredith M, et al. Long-term effect of goal-directed weight management in an atrial fibrillation cohort: The LEGACY study. J Am Coll Cardiol. 2015;65:2159–2169. doi: 10.1016/j.jacc.2015.03.002.

5. Daoud EG, Weiss R, Bahu M, et al. Liraglutide and weight loss for atrial fibrillation burden reduction: The LEAF trial. Heart Rhythm. 2023;20:580–588. doi: 10.1016/j.hrthm.2023.01.015.

6. Rush KL, Burton L, Schaab K, Lukey A. The impact of nurse-led atrial fibrillation clinics on patient and healthcare outcomes: a systematic mixed studies review. Eur J Cardiovasc Nurs. 2019;18:526–533. doi: 10.1177/1474515119845198.

7. Guo Y, Chen Y, Lane DA, et al. Mobile health technology for atrial fibrillation management integrating decision support, education, and patient involvement: mAF App Trial. Am J Med. 2017;130:1388–1396.e6. doi: 10.1016/j.amjmed.2017.04.029.

8. Guo Y, Lane DA, Wang L, et al. Mobile health technology to improve care for patients with atrial fibrillation. Heart Rhythm. 2019;16:1476–1483. doi: 10.1016/j.hrthm.2019.04.018.

9. Turchioe MR, Jimenez V, Isaac S, et al. Review of mobile applications for the detection and management of atrial fibrillation. Heart Rhythm O2. 2020;1:35–43. doi: 10.1016/j.hroo.2020.02.005.

10. Stoyanov SR, Hides L, Kavanagh DJ, Zelenko O, Tjondronegoro D, Mani M. Mobile App Rating Scale: a new tool for assessing the quality of health mobile apps. JMIR Mhealth Uhealth. 2015;3:e27. doi: 10.2196/mhealth.3422.

11. Masterson Creber RM, Maurer MS, Reading M, Hiraldo G, Hickey KT, Iribarren S. Review and analysis of existing mobile phone apps to support heart failure symptom monitoring and self-care management using the Mobile Application Rating Scale (MARS). JMIR Mhealth Uhealth. 2016;4:e74. doi: 10.2196/mhealth.5882.

12. Hayes AF, Krippendorff K. Answering the call for a standard reliability measure for coding data. Commun Methods Meas. 2007;1:77–89. doi: 10.1080/19312450709336664.

13. R Core Team. R: A language and environment for statistical computing. R Foundation for Statistical Computing; 2025. Available from: https://www.R-project.org/.

14. Marent B, Henwood F, Darking M. Development of a mHealth platform for HIV care: a socio-technical co-design approach. JMIR Mhealth Uhealth. 2021;9:e27896. doi: 10.2196/27896.

15. McCabe PJ, Schad S, Hampton A, et al. Perceptions of older adults with atrial fibrillation for engaging with technology to support anticoagulation decision making: a qualitative study. Patient Prefer Adherence. 2020;14:2521–2531. doi: 10.2147/PPA.S278238.

16. Vaportzis E, Giatsi Clausen M, Gow AJ. Older adults’ perceptions of technology and barriers to adopting tablet computers: a focus group study. Front Psychol. 2017;8:1687. doi: 10.3389/fpsyg.2017.01687.

